# Early age at seizure onset is a risk factor for attention deficit hyperactivity disorder in children with epilepsy

**DOI:** 10.1101/2025.03.21.25324382

**Authors:** Iris Caggia, Romain Bouet, Julien Jung, Jean-Philippe Lachaux, Karine Ostrowsky-Coste, Julitta De Bellescize, Vania Herbillon, Marine Thieux

## Abstract

Attention deficit hyperactivity disorder (ADHD) is one of the most common comorbidities in children with epilepsy but the mechanisms underlying their relationship are not clear. To determine whether an earlier age at seizure onset was associated with ADHD independent of other epilepsy-related factors, data from 203 children with epilepsy (6 to 17 years and 11 months; 53% female) were analyzed. The Diagnostic and Statistical Manual of Mental Disorders fifth edition helped define two groups: epilepsy without (n = 96) or with (n = 107) ADHD. The academic repercussions of ADHD were assessed using a composite school impact index. A binary logistic regression model was fitted considering the effects of age at seizure onset, type of epilepsy, frequency of seizures in the last 12 months, and number of antiseizure medication (ASM) on the ADHD diagnosis and their pairwise interactions. Around half of the children (52%) had a comorbid ADHD. The median school impact index was higher in children with epilepsy with vs without ADHD (33 vs 19%, *p* < 0.001). The age at seizure onset had a significant effect on ADHD diagnosis (OR = 0.99, 95% Confidence Interval = 0.96-1.01, *p* = 0.008) independent of other epilepsy-related factors. The risk of ADHD diagnosis decreased by 1% for each additional month of the age at seizure onset. To conclude, the risk of ADHD is higher when seizures begin earlier in life and ADHD exacerbates the adverse impact on school performance, underscoring the imperative for early systematic screening for ADHD from the first seizure.

## 1. Introduction

Childhood epilepsy is associated with many concurrent disorders. Up to 80% of children with epilepsy experience at least one comorbidity, including neurodevelopmental and psychiatric disorders (*e*.*g*. behavioral and emotional disorders, intellectual disability, autism spectrum disorder, unspecified neurodevelopmental delay) [1]. Comorbidities can adversely affect long-term developmental, learning, and psychological outcomes, as well as their quality of life and that of their families [2]. Among these, attention deficit hyperactivity disorder (ADHD) is one of the most common, occurring 2.5 to 5.5 times more frequently in children with epilepsy than in healthy children [3]. However, the mechanisms underlying the relationship between epilepsy and ADHD are not clearly established and there is no consensus regarding the influence of epilepsy-related factors on the presence of ADHD.

Early epilepsy onset is a risk factor for having a psychiatric disorder [4], and is associated with subnormal global cognitive function [5], lower attention performance and impulsivity [6]. A few studies have specifically focused on the impact of the age at seizure onset on the diagnosis of ADHD with contradictory results [7–10]. Epilepsy-related factors (including the age at seizure onset) were not associated with ADHD in a study involving 85 children with active epilepsy [7]. Similarly, there was no age difference at epilepsy onset among 204 children with epilepsy with or without ADHD [8]. In contrast, a significantly lower age at epilepsy diagnosis was observed in 301 children with epilepsy when associated with ADHD [9]. Additionally, a gradual decrease in the risk of ADHD with increasing age at seizure onset has been demonstrated in 206 children with epilepsy [10]. The heterogeneity of previous findings may be attributable to the isolated consideration of the role of the age at seizure onset on a subsequent diagnosis of ADHD, without taking into account the influence of other epilepsy-related factors such as type of epilepsy, frequency of seizures and number of anti-seizure medications.

In the present study, we therefore aimed to determine whether earlier age at seizure onset was associated with the occurrence of ADHD independently of other epilepsy-related factors.

## 2. Methods

### 2.1. Patients

Patients were identified among those of a prospective study conducted between 2015 and 2020. The study protocol was approved by a regional ethics committee (COGNI-AIC-38RC14.374) and recorded on ClinicalTrials.gov (ID: NCT03094793). Children were included during a routine consultation. The inclusion criteria were age between 6 and 18 years, with epilepsy and / or ADHD, having received an electroencephalography (EEG) as part of their diagnosis or follow-up, having given written informed consent to participate in the study (by the child and the legal representative). Patients were excluded from the study if the presence of a disability prevented the completion of the test (severe intellectual disability, severe upper limb motor deficit, blindness). Part of the present study population (122 children, 80 of whom had ADHD) have been described in a previous paper [11]. In the present study, only children with epilepsy diagnosis were included. Patients treated with a psychostimulant at time of the evaluation were excluded.

### 2.2. Material and methods

#### 2.2.1. Demographics

During the interview, demographic information (age, sex assigned at birth, and the socio-economic level [SEL] of the parents according to the national institute of statistics and economical studies [12]) were collected. The effect of epilepsy on education was investigated using a composite “school impact index” calculated using 3 criteria: grade repetition (yes / no), current school level estimated by the parents (very good, good, average, insufficient or very insufficient), and number of ongoing rehabilitations or school interventions (*e*.*g*. speech therapy, psychomotricity, occupational therapy, orthoptics). Each criterion accounts for one-third of the school impact index which ranges from 0 to 100%: a higher score indicates greater educational difficulties (Supplementary materials, Figure S1).

#### 2.2.2. Epilepsy-related data

The diagnosis of epilepsy was made by epileptologists according to the International League Against Epilepsy classification [13]. The following clinical data concerning epilepsy were collected: the age at seizure onset (in months), the type of epilepsy (generalized, focal, or unknown), the frequency of seizures in the last 12 months (no seizure, 1 to 12, 13 to 52, or at least 1 per day), the number of anti-seizure medications (ASM) at the time of the consultation (no treatment, 1, 2 or more) and the syndrome classification (generalized epilepsies: childhood absence epilepsy, juvenile absence epilepsy, juvenile myoclonic epilepsy, other idiopathic generalized epilepsy, generalized symptomatic epilepsy, generalized epilepsy of unknown cause, and epilepsy with eyelid myoclonia; focal epilepsies: focal symptomatic epilepsy, self-limited epilepsy with centrotemporal spikes, self-limited epilepsy with autonomic seizures, and focal epilepsy of unknown cause).

#### 2.2.3. ADHD diagnosis

A systematic screening of ADHD, with complaints or not by the patient, was performed during the clinical evaluation by an experienced pediatric epileptologist and a neuropsychologist. The ADHD diagnosis (yes / no) and subtype (inattention, hyperactivity-impulsivity, or combined) was based on the Diagnostic and Statistical Manual of Mental Disorders fifth edition (DSM-V) criteria [14]. The severity of ADHD was assessed using the ADHD Rating-Scale IV (ADHD-RS IV) [15], based on the DSM-V criteria and rated on a Likert-scale ranging from 1 to 4 points. Two separate scores are calculated to differentiate symptoms of inattention from those of hyperactivity-impulsivity and then combined to obtain a total score. The higher the scores, the more severe the disorder.

### 2.3. Statistical analysis

Two groups of children were formed based on the ADHD diagnosis: the epilepsy without ADHD group, and the epilepsy with ADHD group. The R software (version 4.4.1) was used for statistical analyses [16]. P-values less than 0.05 were considered statistically significant. Missing data are reported in the tables and were not included in the statistical analyses. Firstly, a descriptive analysis was conducted considering the two groups. Quantitative data are presented as median (range), and qualitative data as number (percentage). Subsequently, comparisons between the two groups regarding demographic variables were conducted using Wilcoxon tests for qualitative data (age, school impact index), and chi-square tests for quantitative data (sex and SEL). Finally, the influence of epilepsy-related factors on the diagnosis of ADHD was evaluated using a binary logistic regression model (glm function, R-stats 4.4.1). The variable to be explained was the diagnosis of ADHD (yes / no), the explanatory variables were the epilepsy-related factors (age at seizure onset, type of epilepsy, frequency of seizures in the last 12 months, and number of ASM at evaluation). The effects of the explanatory variables and their pairwise interactions were tested applying a Wald test on the model (Anova function, R-car 3.1.2).

## 3. Results

Among the 203 children included, three had an epilepsy of unknown type. These children were 145, 84 and 72 months old at seizure onset, and had a comorbid diagnosis of ADHD (one combined form and two inattention form). These three were excluded from the statistical model because epilepsy with unknown type was under-represented.

### 3.1. Descriptive analysis

The 203 children (53% female) were aged 6 years to 17 years and 11 months (median age: 10 years and 3 months). The median school impact index was 28% (range: 0%-100%; Table 1). The median age at seizure onset was 72 months, half of the patients had a generalized epilepsy, the majority had 12 seizure or less during the past year and only one ASM. Epilepsy-related characteristics are described in Table 2 (for the syndromes see Supplementary materials, Table S1). Regarding the ADHD diagnosis, 52% met the criteria for the diagnosis of ADHD, 8% with the hyperactive-impulsivity form, 61% with the inattention form, and 31% with the combined form. For children with ADHD, the median ADHD-RS inattention score was 18 (range: 5-27), 12 (range: 0-24) for hyperactivity-impulsivity score, and 29 (range: 12-46) for the total score.

**Table 1.** Demographic characteristics of children with epilepsy without and with ADHD.

|  | Total<br>population n =<br>203 | ADHD |  | p-value |
| --- | --- | --- | --- | --- |
|  |  | Without<br>n = 96 | With<br>n = 107 |  |
| Age at examination (months) | 124 (72-215) | 131 (72-215) | 118 (72-209) | 0.257 |
| Sex (F) | 107 (52.7%) | 57 (59.4%) | 50 (46.7%) | 0.097 |
| Socio-economic level of the parents |  |  |  | 0.142 |
| Farmers | 0 (0.0%) | 0 (0.0%) | 0 (0.0%) |  |
| Craftsmen, Traders, and Business Owners | 26 (12.8%) | 9 (9.4%) | 17 (15.9%) |  |
| Managers and Professionals | 51 (25.1%) | 24 (25.0%) | 27 (25.2%) |  |
| Intermediate Professions | 23 (11.3%) | 16 (16.7%) | 7 (6.5%) |  |
| Employees | 79 (38.9%) | 39 (40.6%) | 40 (37.4%) |  |
| Manual Workers | 15 (7.4%) | 5 (5.2%) | 10 (9.3%) |  |
| Retirees | 0 (0.0%) | 0 (0.0%) | 0 (0.0%) |  |
| Others or without professional activity | 9 (4.4%) | 3 (3.1%) | 6 (5.6%) |  |
| School impact index | 28 (0-100) | 19 (0-83) | 33 (0-100) | <0.001 |

Results are presented as median (range) or n (%). ADHD: attention deficit hyperactivity disorder; ASM: anti-seizure medication. P-value: Wilcoxon test (qualitative data) or Chi-square test (quantitative data).

**Table 2.**
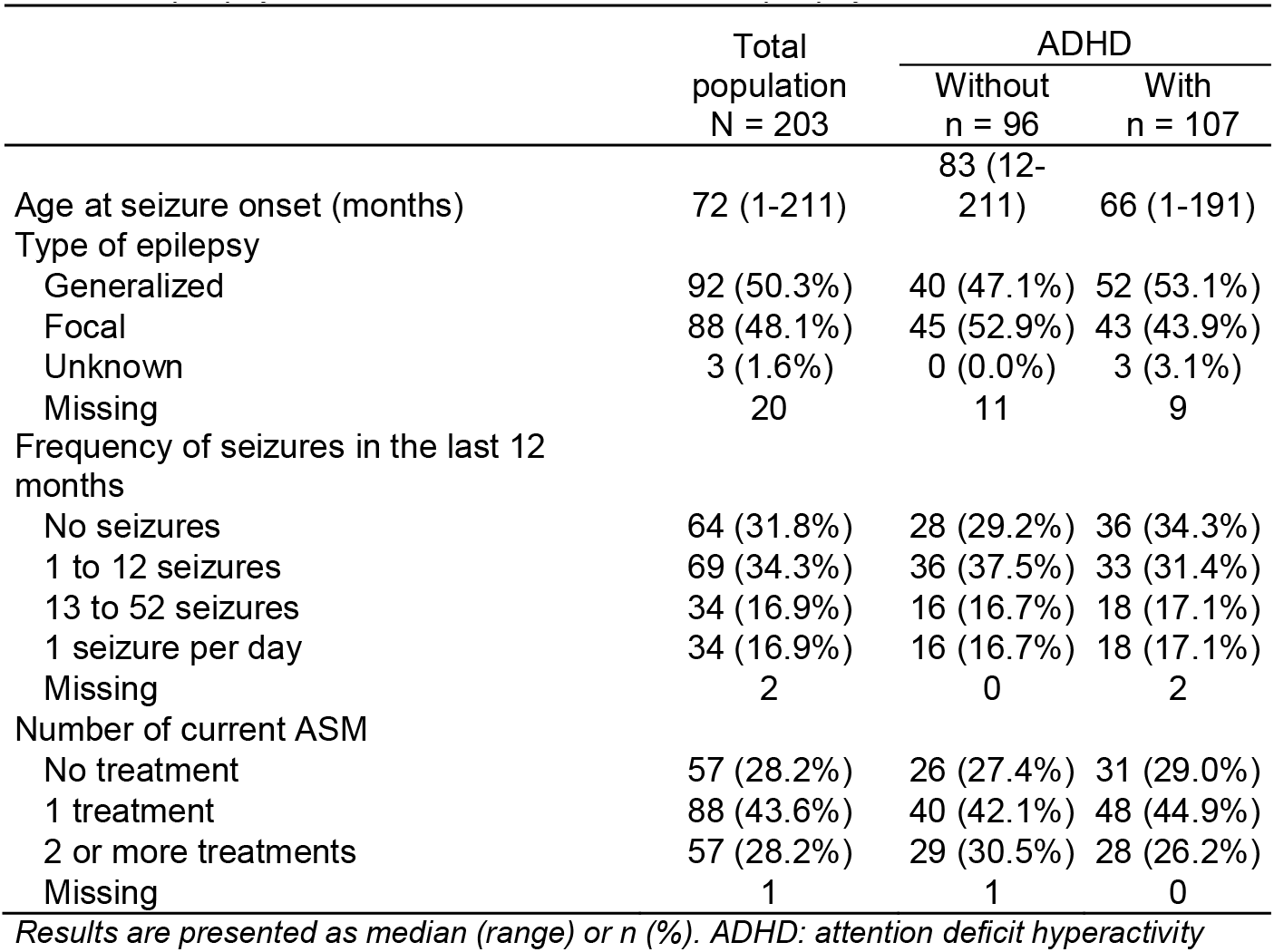
Epilepsy-related factors of children with epilepsy without and with ADHD.

|  | Total<br>population<br>N = 203 | ADHD |  |
| --- | --- | --- | --- |
|  |  | Without<br>n = 96 | With<br>n = 107 |
| Age at seizure onset (months) | 72 (1-211) | 83 (12-211) | 66 (1-191) |
| Type of epilepsy |  |  |  |
| Generalized | 92 (50.3%) | 40 (47.1%) | 52 (53.1%) |
| Focal | 88 (48.1%) | 45 (52.9%) | 43 (43.9%) |
| Unknown | 3 (1.6%) | 0 (0.0%) | 3 (3.1%) |
| Missing | 20 | 11 | 9 |
| Frequency of seizures in the last 12 months |  |  |  |
| No seizures | 64 (31.8%) | 28 (29.2%) | 36 (34.3%) |
| 1 to 12 seizures | 69 (34.3%) | 36 (37.5%) | 33 (31.4%) |
| 13 to 52 seizures | 34 (16.9%) | 16 (16.7%) | 18 (17.1%) |
| 1 seizure per day | 34 (16.9%) | 16 (16.7%) | 18 (17.1%) |
| Missing | 2 | 0 | 2 |
| Number of current ASM |  |  |  |
| No treatment | 57 (28.2%) | 26 (27.4%) | 31 (29.0%) |
| 1 treatment | 88 (43.6%) | 40 (42.1%) | 48 (44.9%) |
| 2 or more treatments | 57 (28.2%) | 29 (30.5%) | 28 (26.2%) |
| Missing | 1 | 1 | 0 |

Results are presented as median (range) or n (%). ADHD: attention deficit hyperactivity

### 3.2. Between group comparisons

Comparison of children with epilepsy with and without ADHD revealed no significant difference regarding age, sex, or SEL (*p* > 0.05, Table 1). The median school impact index was significantly higher in the group of children with epilepsy with ADHD compared to the group with epilepsy without ADHD (33% vs 19%, *p* < 0.001, Table 1, Figure 1).

**Figure 1.**
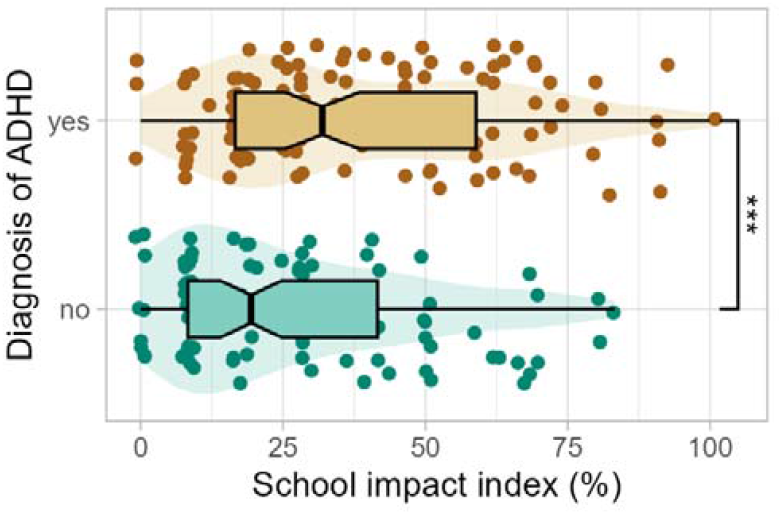
School Impact Index according to attention deficit hyperactivity disorder (ADHD) diagnosis. School Impact Index of children with epilepsy with ADHD (orange) and without ADHD (green). Each point represents the school impact index for each child with epilepsy according to the diagnosis of ADHD. The central line of boxplots corresponds to the median, the left and right edges correspond to the first and third quartiles, and the whiskers display the ranges. The violin plots describe the distributions. Difference between groups is represented by stars: ***: p <0.001.

### 3.3. Association between epilepsy and ADHD

The binary logistic linear regression including all epilepsy-related variables and their 2 by 2 interactions showed that age at seizure onset had a significant effect on ADHD diagnosis (*p* = 0.008). The risk of ADHD diagnosis decreased by 1% for each additional month of the age at seizure onset (odds ratio, OR = 0.99, 95% Confidence Interval (CI) = 0.96 – 1.01, Figure 2). The median age at seizure onset was lower in the group with epilepsy with ADHD than in the group with epilepsy without ADHD (66 vs 83 months, Figure 2). The type of epilepsy, frequency of seizures in the last 12 months, and number of ASM did not have a significant effect on the diagnosis of ADHD according to the model, and their distributions were similar between the two groups (Supplementary materials, Figure S2 and Table S2).

**Figure 2.**
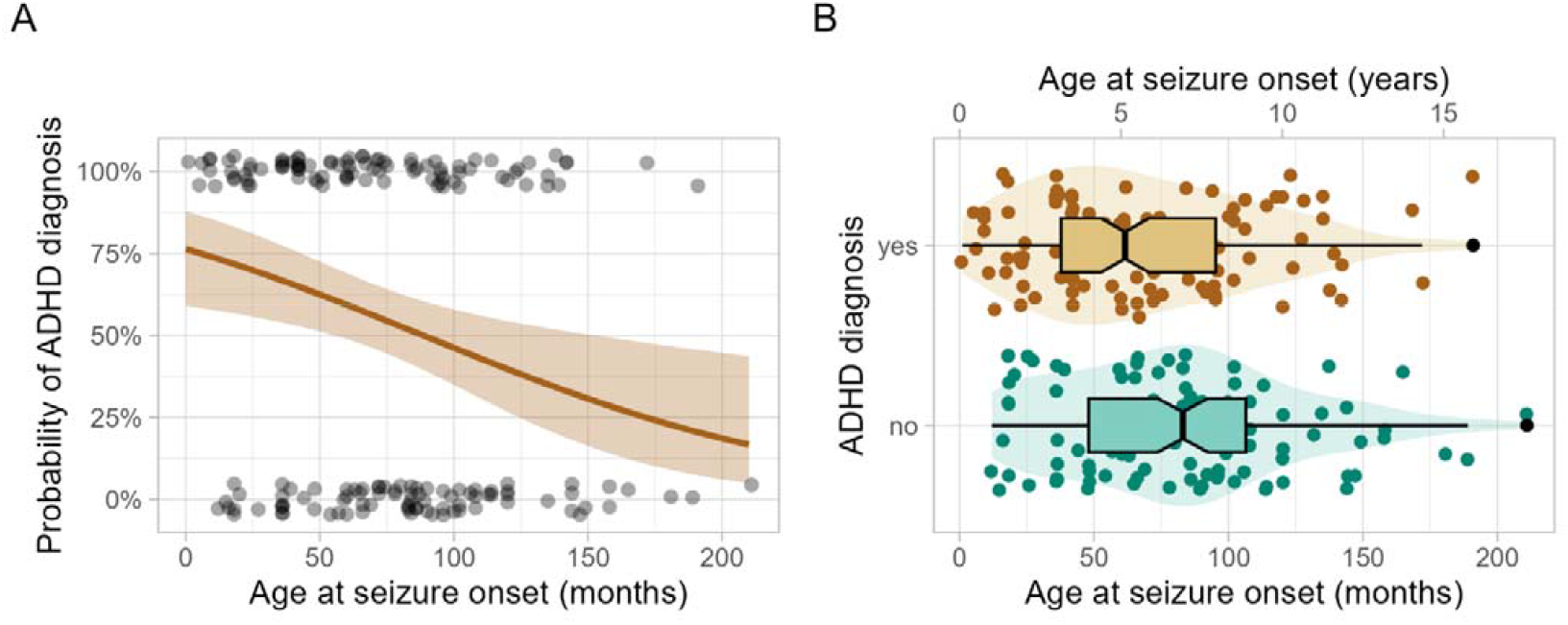
ADHD diagnosis according to the age at seizure onset in children with epilepsy. **(A)** Predicted probability of ADHD diagnosis according to age at seizure onset (dark orange). The 95% confidence interval is represented in light orange. Each point represents the age at seizure onset of each child with (at the top) or without ADHD (at the bottom). **(B)** Age at seizure onset in children with epilepsy without (green) and with ADHD (orange). Each point represents the age at seizure onset for each child with epilepsy according to the diagnosis of ADHD. The central line of boxplots corresponds to the median, the left and right edges correspond to the first and third quartiles, and the whiskers display the ranges. The violin plots describe the distributions.

## 4. Discussion

In addition to highlighting that comorbid ADHD is associated with a higher impact on school performance in children with epilepsy, the present study also demonstrates that a younger age at seizure onset is associated with a higher risk of developing a comorbid ADHD, independent of other epilepsy-related factors.

Our cohort reflect what is typically found in pediatric epilepsy studies: half of the patients had an associated ADHD [2], with a predominance of the inattentive form [9,10,17], and an equal sex distribution within the group with epilepsy with ADHD [3,17]. Children with epilepsy with ADHD had a higher school impact index than those without, corroborating that the combination of epilepsy and ADHD is associated with a negative impact on children’s academic performance in school [18].

The relationship between epilepsy and ADHD in children could be explained by several hypotheses involving epilepsy-related factors. First, while ADHD symptoms could be understood as side-effects of ASM, no significant effect of ASM on ADHD was found in the present study, consistent with previous studies [9,10]. Second, there may be a direct causal relationship between epilepsy and ADHD, with factors intrinsic to epilepsy acting as the noxious agent and influencing the expression, severity and characteristics of ADHD [19]. For instance, attention difficulties has been shown to be more common among generalized epilepsies [2] but our analysis did not reveal any effect of epilepsy type on ADHD diagnosis. In addition, the frequency of seizures had no effect on the ADHD diagnosis, consistent with a previous study [10]. One might also question the influence of epileptic discharges on brain activity across development. Interictal epileptiform discharges have been related to cognitive and sustained attention impairment [6][20], especially when interfering with (slow-wave) sleep [21], as observed in children with electrical status epilepticus in slow-wave sleep [22]. Beyond interictal discharges, epileptic activity could interfere with brain maturation, especially when occurring within a critical developmental window, thus impacting maturational processes (*e*.*g*. cellular proliferation, migration, neuronal differentiation, synaptic integration), as well as neuronal activity and plasticity [23]. According to the maturity of the brain at the onset of the disease, epileptic activity can thus durably impact cognition to varying degrees [6,23,24]. Interestingly, indicators for poorer or delayed brain maturation have been observed in both benign childhood epilepsy with centrotemporal spikes [25] and in ADHD without epilepsy (specifically in (pre)frontal regions) [26,27]. In addition, and consistent with previous studies [9–11][28], our results suggest that the early development of epilepsy could influence brain maturation processes with lasting impacts on attention functioning. An increase in the age at seizure onset was associated with a decreased risk of an ADHD diagnosis, regardless of the type of epilepsy, the frequency of seizures or the number of ASM. While the existence of a critical period during which epileptic activity affects attention networks needs to be further clarified, a previous study showed that the risk of an ADHD diagnosis was greater than 50% when seizures begun during a critical 5-year period [5]. Finally, other factors which are not considered herein could also play a role in the co-occurrence of epilepsy and ADHD, such as the interaction between genetic [29] and environmental factors (*e*.*g*. toxins, low birth weight, advanced maternal age, and psychosocial deprivation) [2] or an underlying brain damage / dysfunction from various etiologies [19,20]. Noticeably, ADHD symptoms can also precede the first seizure, suggesting an influence of pre-existing neurobiological factors, independent of epilepsy-related factors.

The present study has certain limitations. To begin with, our cohort presents a variety of epileptic syndromes that were not considered in the analyses due to an insufficient number of patients in each subgroup. Further studies are required to investigate the relationship between ADHD and epilepsy-related characteristics within specific syndromes. Second, the hospital unit from which the data were obtained performs pre-surgical assessment of epilepsy. Therefore, the proportion of focal drug-resistant epilepsies is higher than that of generalized drug-resistant epilepsies, which may increase the incidence of attention impairment in focal epilepsy in our cohort. Furthermore, interictal epileptiform discharges were not considered and only seizures during the last year were taken into account. In addition, it would be valuable to examine the impact of the age at seizure onset on the severity of ADHD. However, quantifying severity is challenging using the DSM-V criteria, as symptoms tend to diminish in intensity with age, despite the persistence of ADHD. The total score of the ADHD-RS is not an ideal measure either, as two children who have the same score may have disorders of different intensities (*e*.*g*. one child may have intense inattention, while the other has some moderate hyperactivity-impulsivity and inattention). One solution may lie in neuropsychological attention tests, which may provide a more nuanced assessment of severity. It is also worth considering whether the ADHD diagnosis depends on the active duration of the disease rather than on the age at seizure onset, given the strong connection between these two variables. A prospective longitudinal study could greatly help to disentangle these factors by following children admitted for suspected epilepsy and enabling the precise determination of the timing of both epilepsy and ADHD diagnoses. Screening for ADHD at seizure onset could determine whether the disorder precedes or co-occurs with epilepsy.

## 5. Conclusion

In conclusion, the risk of ADHD diagnosis is significantly higher when seizures begin earlier in life, independent of other epilepsy-related factors, and decreases by 1% for each additional month of the age at seizure onset. Additionally, ADHD exacerbates the adverse impact on academic success in children with epilepsy, underscoring the imperative for early systematic screening for ADHD from the first seizure, to promote the implementation of suitable clinical care, thereby reducing the impact of this prevalent comorbidity.

## Data Availability

The data supporting the results of this study are not publicly available due to ethical and privacy restrictions. The data may be available on request from the corresponding author, with the additional and documented consent of the patients and legal representatives involved in this study.

## Author contributions

**Iris Caggia**: Software, Formal analysis, Data curation, Writing – original draft, Visualization. **Romain Bouet**: Methodology, Software, Validation, Writing – review and editing. **Julien Jung:** Writing – review and editing. **Jean-Philippe Lachaux**: Resources, Writing – review and editing. **Karine Ostrowsky-Coste**: Resources, Writing – review and editing. **Julitta De Bellescize:** Conceptualization, Investigation, Resources, Writing – review and editing. **Vania Herbillon**: Conceptualization, Investigation, Data curation, Writing – review and editing, Supervision, Project administration. **Marine Thieux**: Conceptualization, Methodology, Validation, Writing – review and editing, Supervision.

## Supplementary Materials

**Figure S1.**
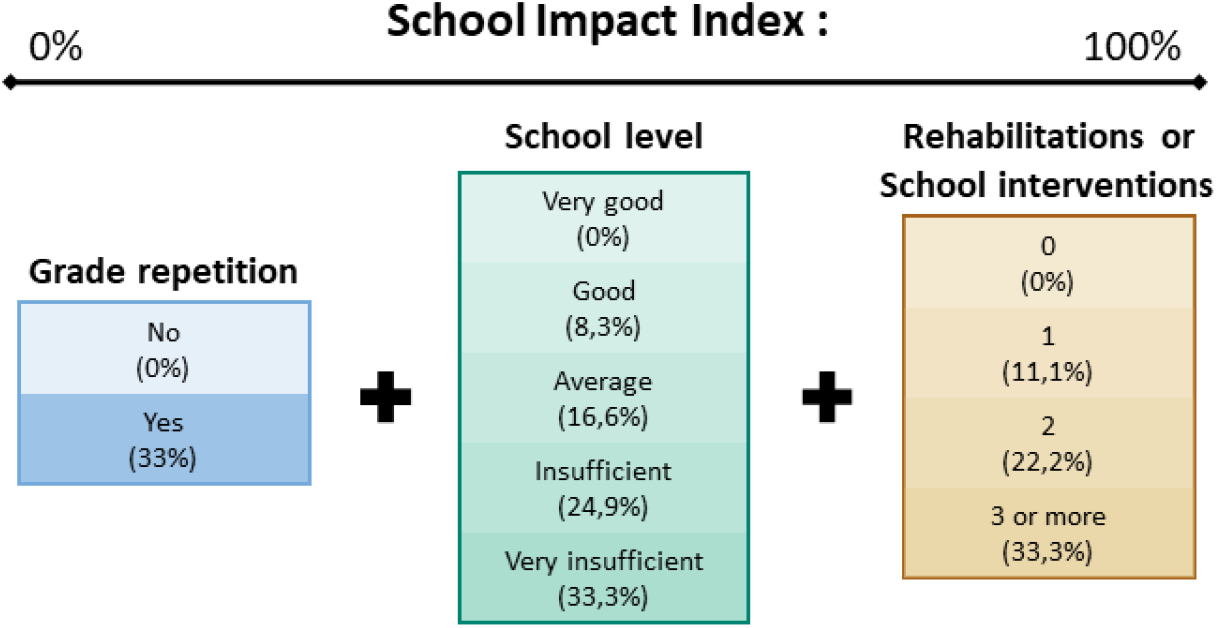
School Impact Index. The School Impact Index assesses the repercussions of attention deficit hyperactivity disorder on scholar achievement. Ranging from 0% to 100%, it is calculated according to three criteria: presence or absence of grade repetition, current school level estimated by the parents and number of ongoing rehabilitations or school interventions. Each criterion account for one third of the School Impact Index, the percentages attributed to them are summed. The higher the School Impact Index, the greater the academic difficulties.

**Figure S2.**
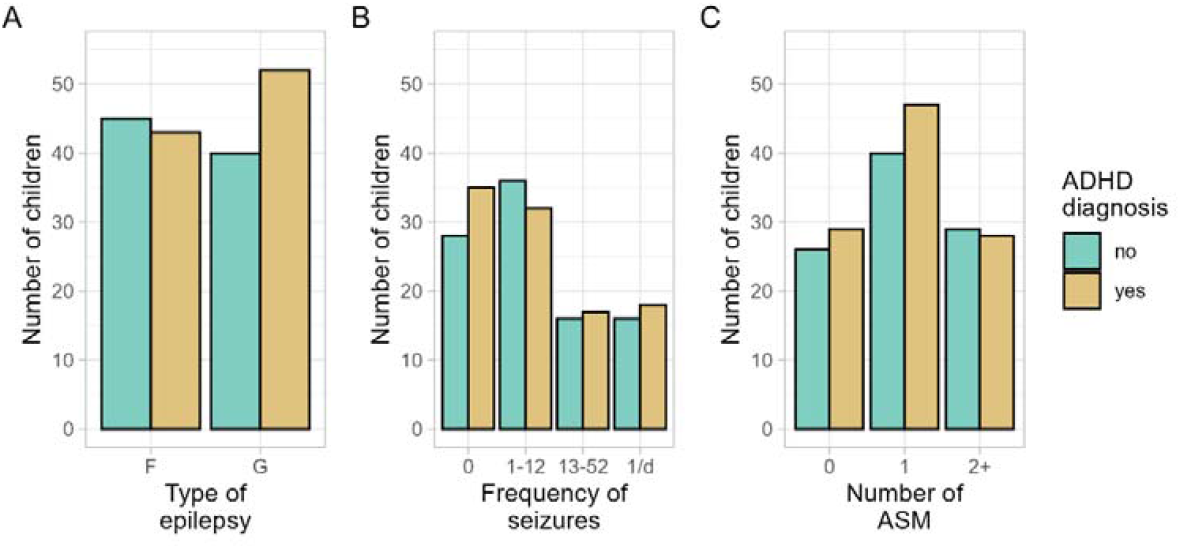
Epilepsy-related factors in children with epilepsy according to ADHD diagnosis. Epilepsy-related factors in children with epilepsy without (green) and with ADHD (orange). **(A)** Type of epilepsy: focal (F) or generalized (G). **(B)** Frequency of seizures in the last 12 months: No seizure (0), 1 to 12 seizures (1-12), 13 to 52 seizures (13-52), and 1 seizure per day or more (1/d). **(C)** Number of current anti-seizure medication (ASM). ADHD: attention deficit hyperactivity disorder.

**Table S1.** Epileptic syndromes of children with epilepsy without and with ADHD.

|  | Total population<br>N = 203 | ADHD |  |
| --- | --- | --- | --- |
|  |  | Without<br>n = 96 | With<br>n = 107 |
| Focal symptomatic epilepsy | 13 (7.1%) | 5 (5.9%) | 8 (8.2%) |
| Focal epilepsy of unknown cause | 22 (12.0%) | 13 (15.3%) | 9 (9.2%) |
| Self-limited epilepsy with<br>centrotemporal spikes | 38 (20.8%) | 20 (23.5%) | 18 (18.4%) |
| Self-limited epilepsy with autonomic<br>seizures | 14 (7.7%) | 6 (7.1%) | 8 (8.2%) |
| Childhood absence epilepsy | 29 (15.8%) | 14 (16.5%) | 15 (15.3%) |
| Juvenile absence epilepsy | 6 (3.3%) | 3 (3.5%) | 3 (3.1%) |
| Juvenile myoclonic epilepsy | 1 (0.5%) | 1 (1.2%) | 0 (0.0%) |
| Other idiopathic generalized epilepsy | 30 (16.4%) | 11 (12.9%) | 19 (19.4%) |
| Generalized symptomatic epilepsy | 4 (2.2%) | 3 (3.5%) | 1 (1.0%) |
| Generalized epilepsy of unknown<br>cause | 12 (6.6%) | 4 (4.7%) | 8 (8.2%) |
| Epilepsy with eyelid myoclonia | 11 (6.0%) | 5 (5.9%) | 6 (6.1%) |
| Epilepsy of unknown type | 3 (1.6%) | 0 (0.0%) | 3 (3.1%) |
| Missing | 20 | 11 | 9 |

*Results are presented as n (%). ADHD: attention deficit hyperactivity disorder*

**Table S2.** Logistic (binomial) regression (without interactions).

|  | Odds ratio | 95% Confidence Intervals | p-value |
| --- | --- | --- | --- |
| Age at seizure onset, months | 0.99 | 0.98-1.00 | 0.010 |
| Epilepsy type, generalized (vs focal) | 1.53 | 0.80-2.96 | 0.200 |
| Seizure frequency (past year) |  |  |  |
| 1 to 12 (vs none) | 0.86 | 0.40-1.83 | 0.690 |
| 13 to 52 (vs none) | 1.19 | 0.45-3.20 | 0.722 |
| 1 / day (vs none) | 0.77 | 0.29-2.06 | 0.602 |
| ASM |  |  |  |
| 1 (vs 0) | 0.79 | 0.37-1.67 | 0.537 |
| 2+ (vs 0) | 0.78 | 0.31-1.94 | 0.595 |

